# A snapshot of the UK blood donor plasma virome: a retrospective cross-sectional cohort study

**DOI:** 10.64898/2026.05.20.26353651

**Authors:** Kai Kean, Richard M. Mayne, Kaitlin Reid, Shannah Secret, Belinda K. Singleton, Rebecca Rockett, Piya Rajendra, Heli Harvala, Judith Breuer, M. Azim Ansari, Katrina Lythgoe, Peter Simmonds, Tanya Golubchik

## Abstract

Estimates of population prevalence and genetic diversity of bloodborne viruses in healthy humans are essential to support population-scale monitoring for transfusion transmission risk. In the UK and globally, blood donations are routinely screened for a limited number of high-consequence pathogens, but the full composition of the plasma virome remains to be characterised. Using a novel quantitative targeted metagenomics sequencing approach, we analysed previously unscreened plasma donations collected by NHS Blood and Transplant in England for all major pathogenic and known commensal human bloodborne viruses, and quantified their viral burden. Here we show that in a representative sample of 5,064 UK blood donors in pools of 24 collected over a one-month period, the virome was dominated by a small number of largely persistent species, representing ∼11% (12/106) of previously identified human bloodborne viruses. Anelloviruses (TTV, TTMV and TTMDV) was detected in 89.0% of pools, albeit at low read count inconsistent with measured anellovirus viral loads. In contrast, human pegivirus type 1 (HPgV-1), had estimated population prevalence of 3.7% (95% CI 3.0—4.4%), with high read count and complete genome recovery in around one half of positive pools, consistent with high titre in plasma. Estimated prevalences for less common detections included one species of gemykibovirus (0.12%), hepatitis C virus (genotype 1a, 0.04%) and various polyomaviruses and herpesviruses between 0.04% (parvovirus 4, BK polyomavirus) and 0.41% (human herpesvirus 6). Phylogenetic analyses revealed mixed TTV, TTMV and TTMDV populations and almost exclusively genotype 2 HPgV-1, consistent with known genotype distributions in Europe. Our results provide a baseline for describing the healthy plasma virome in UK blood donors.

## 1 Introduction

The NHS Blood and Transplant (NHSBT) blood service in England collects 1.8 M blood donations per year^1^ from a pool of almost 800,000 donors^2^. Proven transfusion-transmitted infections (TTIs) are exceptionally rare in England and elsewhere in the UK, with just 14 confirmed transmissions of major pathogens (HIV-1, HBV and HCV) reported between 2015-2024 (Narayan & SHOT Steering Group, 2024). Routine testing targets only a limited number of high-consequence viruses, and the wider composition of the human blood-borne virome remains to be characterised.

The human blood virome is known to vary along a number of demographic lines including gender, age, ethnicity and geographical location, but infections with anelloviruses, human pegivirus type 1 (HPgV-1), polyomaviruses and herpesviruses are common throughout the world (Moustafa et al., 2017; Pyöriä et al., 2024; Rascovan et al., 2016; Waldvogel-Abramowski et al., 2019; Wylie et al.,2014). Anelloviruses in particular are very frequently detected in the blood of healthy people (Sabbaghian et al., 2024) and are generally considered entirely non-pathogenic. Their study is regardless valuable, as changes in the titres of anellovirus torque teno virus (TTV) were recently demonstrated to be predictive for the effectiveness of immunosuppressive chemotherapy, systemic infections by other pathogens and in transplant graft rejection (Görzer et al., 2023; Widder et al., 2022). Viral loads in blood can be considered an indicator of the extent of systemic viral replication balanced by the effectiveness of the immune system in clearing cell-free virus. Methods that both sequence and quantify viruses in plasma have recently been developed (Bonsall et al., 2020; Lin et al., 2024; Lythgoe et al., 2021).

In a blood donation context in the UK, current molecular and serological testing is targeted towards for 5 viruses (HIV-1 and -2, HBV, HCV, HEV, and additional testing for hepatitis A virus [HAV] and parvovirus B19 [B19V] in plasmapheresis donors), and discretionary testing for four others, human cytomegalovirus (HCMV, human herpesvirus 5), human T-cell lymphotropic virus -1 and -2, and West Nile virus, although the possibility of emergence of novel pathogens is the subject of ongoing horizon scanning and review by the blood services (McClure et al., 2024; Neuberger et al., 2023; Rajendra et al., 2026). For that purpose and to better understand the range of viruses infecting healthy blood donors who are at least partly representative of the wider population, we have implemented and further developed high-throughput sequencing (HTS) technologies that provide an exceptionally broad screening capability for viruses in blood. Assay development has overcome the technical challenges in implementing new technologies at scale to provide an extensive cross-sectional view of human virome composition. The targeted metagenomics approach we developed uses a bespoke hybridisation probe set to screen plasma from a large cohort of healthy UK blood donors, equivalent to approximately one day of national blood collection donations or around 0.6% of the 2022 donor pool, and presents a range of quantitative and phylogenetic analyses to describe characteristic components of the donor plasma virome.

## 2 Methods

### 2.1 Sample provenance

Validation of assay sensitivities and target quantitation was performed on a range of externally validated, regulator-approved control samples of eight blood-borne viruses that represent a range of genome compositions and organisations (Table 1). Controls were purchased as reagents from the National Institute for Biological Standards and Control (NIBSC, https://nibsc.org, part of the Medicines and Healthcare products Regulatory Agency, MHRA) and Paul Erhlich Instiute (PEI). These included six viruses from quantified international standards (NIBSC reagents 11/242, 22/120, 14/212; PEI 7657/12): human parechovirus (HPeV) type 3, Epstein-Barr virus (EBV, human herpesvirus 4), HCMV, BK polyoma virus (BKV), hepatitis B virus (HBV) and hepatitis D virus (HDV). All standard control samples were run as a dilution series of 1:1, 1:10, 1:100, 1:1000. Additionally, a range of clinical plasma samples from NHSBT known to be positive for hepatitis E virus (HEV) and hepatitis C virus (HCV) (*n* = 20 each) were analysed and separately quantified in-house using quantitative PCR (qPCR).

**Table 1:** Manufacturer/provider details for all sequenced control and clinical samples. cp/mL: copies per millilitre; IU/mL: International Units per millilitre, *n*: number of samples.

| Sample | Source | Dataset | Virus | Genome Type | n | Log10(Viral load) |
| --- | --- | --- | --- | --- | --- | --- |
| Multiplex reagent 11/242 (Mee et al., 2016) | NIBSC | Control | HPeV | RNA | 3 | 4.07 - 7.07 cp/mL |
|  |  |  | HCMV | DNA | 3 | 1.66 - 4.66 cp/mL<br>2.07 - 5.07 IU/mL |
|  |  |  | EBV | DNA | 3 | 0.88 - 3.88 cp/mL<br>0.81 - 3.81 IU/mL |
| BKV 14/212 (Govind et al., 2017) | NIBSC | Control | BKV | DNA | 3 | 4.20 - 7.20 IU/mL |
| HDV 7657/12 (Pyne et al., 2017) | PEI | Control | HDV | RNA | 3 | 2.76 - 5.76 IU/mL |
| HBV 22/120 (Heermann et al., 1999) | NIBSC | Control | HBV | RNA | 3 | 1.65 - 4.65 IU/mL |
| HCV-positive plasma | NHSBT | Control | HCV | RNA | 20 | 3.63 - 6.34 IU/mL |
| HEV-positive plasma | NHSBT | Control | HEV | RNA | 20 | 1.63 - 4.77 IU/mL |
| Donor pooled plasma | NHSBT | Experimental | - | - | 211 | - |

211 unscreened plasma pools from UK blood donors, each representing 24 donors, were sourced from NHS Blood and Transplant (NHSBT), all from Southern England in August of 2022, of which 20% were from London. Demographics for the entire plasma pool cohort are included in S.I. 2.1.

### 2.2 Nucleic acid extraction

Pools were extracted with the Thermo Scientific KingFisher Apex Automated Extraction System using Zymo Quick DNA/RNA Viral MagBead kit R2141 (Cambridge Bioscience). Extraction volume was 200 µL and elution volume was 35 µL, according to manufacturer instructions.

### 2.3 PCR

#### 2.3.1 HCV qPCR

Viral loads for HCV-positive clinical plasma were measured using RT-qPCR assays using the TRUPCR HCV Viral Load Kit (TRUPCR, Manchester, UK) according to manufacturer instructions.

#### 2.3.2 HEV qPCR

Virus RNA was detected by RT-qPCR assay as previously described, using primers modified by a 5⍰ flap region and alternative 5⍰-reporter and 3⍰-quencher dyes (MAF, TAMRA) (McClure et al., 2024). Viral load measurements were calibrated to IU/mL using external standards from the National Institute for Biological Standards and Control (NIBSC).

#### 2.3.3 Anellovirus qPCR

Assays were calibrated by reference to a medium virus load plasma sample from a single anonymous blood donor (NHSBT). DNA was extracted from 200 µL of plasma using the QIAamp DNA Blood Mini Kit (Qiagen) and eluted in 50 µL nuclease-free distilled water. Semi-quantitation of the TTV viral loads in 80 of the plasma pools was performed using a modification of the PCR assay described by Maggi *et al*. (Maggi et al., 2003). Reactions were performed on a QuantStudio 3 real time PCR machine, with SYBR Select Master Mix (Thermo Fisher Scientific) and 400 nM primers (no probe). Cycling was 95°C 5 min, with 45 cycles of 95°C 15 sec, 58°C 1 min, and included a dissociation curve. Limiting dilution PCR (with 12 to 24 replicates at 2- to 4-fold serial dilutions of input DNA) was used to quantify detectable TTV copies in the control sample; based on the Poisson distribution at 1/512, 1/1024 and 1/2048 dilutions, an estimate of 51400 copies/ml (log_10_ viral load 4.71) in the original sample was made.

For viral load quantitation of the plasma pools, PCR was performed in reaction volumes of 20 µL, including 4 µL of each plasma pool DNA (extracted from 200 µL plasma and eluted in a volume of 35 µL). A standard curve was generated using 4-fold serial dilutions of the control DNA and run alongside the plasma pool reactions.

#### 2.3.4 Genus-specific amplification of anelloviruses

Nested or semi-nested PCRs for TTV (*Alphatorquevirus*), TTMV (*Betatorquevirus*) and TTMDV (*Gammatorquevirus*) were based on those described by Ninomiya et al. (Ninomiya et al., 2008). Primary reactions (25 µL) contained 5 µL DNA, 0.625 units of GoTaq G2 DNA polymerase (Promega), 1x GoTaq DNA polymerase reaction buffer, 1.5 mM MgCl_2_, 200 nM primers and 0.2 mM dNTPs. PCR cycling was 95°C for 2 min, followed by 35 cycles of 95°C 30 sec, 58°C 30 sec, 72°C 30 sec with a final extension of 72°C 7 min. Genus-specific secondary rounds of PCR (20 µL) used 2 µL of primary product diluted 1/10 and were performed on a QuantStudio 3 real time PCR machine, with SYBR Select Master Mix (Thermo Fisher Scientific). Cycling was 95°C 5 min, with 45 cycles of 95°C 15 sec, 58°C 1 min, and included a dissociation curve.

#### 2.3.5 HPeV nested PCR

Viral RNA was prepared as previously described. The 5’ UTR region was amplified according to (Harvala et al., 2008), with the exception of the addition of a two-minute denaturation step at 95°C at the beginning of the reaction. Outer sense and antisense primers used were5⍰GGGTGGCAGATGGCGTGCCATAA (253) and 5⍰CCTRCGGGTACCTTCTGGGCATCC (583), and inner sense and antisense primers were 5⍰YCACACAGCCATCCTCTAGTAAG (313) and 5⍰GTGGGCCTTACAACTAGTGTTTG (556).

### 2.4 Library preparation and sequencing

Complementary DNA (cDNA) synthesis was performed prior to library preparation, using SuperScript IV VILO Master Mix (Thermo Fisher Scientific, MA, USA) for first strand, and NEBNext Second Strand Synthesis Kit (New England Biolabs, Massachusetts, USA), both according to manufacturer instructions.

Sequencing libraries were then prepared using the Twist Library Preparation EF 2.0 kit, according to a modified half-volume protocol based on the Twist library preparation guidelines. Briefly, DNA fragmentation, end repair, and A-tailing were performed using the Twist Library Preparation Enzymatic Fragmentation Kit at half volumes, with 5.0 µL of cDNA combined with 20 µL of master mix. 2.5 µL of Twist universal adapters were then ligated to the A-tailed DNA fragments using the Twist ligation mix, followed by incubation at 20^⍰^C for 15min. Purification beads were added to each well and samples were washed twice with 150 µL of 80% ethanol, air dried on a magnetic plate, and resuspended in 8.5 µL of water or buffer EB. From this, 7.5 µL of ligated and indexed libraries were transferred to a 96-well plate.

To amplify the libraries, 5.0 µL of Twist UDI primers and 12.5 µL of Equinox Library Amp Mix were added to each well. The libraries were subjected to 16 cycles of PCR amplification and purified post-amplification using AMPure XP DNA purification beads (Beckman Coulter, California, USA). Libraries were quantified with the Thermo Fisher Qubit dsDNA Broad Range Quantitation Assay.

For hybridisation, a 3.0 µL aliquot of each amplified library was pooled, pulse-spun, and dried without heat in a rotary vacuum concentrator. Libraries were resuspended with Twist Universal Blockers and blocker solution, heated to 95^⍰^C, and combined with probe solutions. After adding 15 µL of hybridisation enhancer, the mixture was sealed and incubated at 70^⍰^C for 16h.

Post-capture pools were incubated with streptavidin beads, washed as per protocol, and resuspended in 22.5 µL of water. Illumina amplification primers and Equinox Library Amp Mix were added to the slurry, and libraries underwent 9 cycles of PCR amplification. Following DNA purification and 80% ethanol washes, a 15 µL aliquot of enriched library was recovered and quantified with the Thermo Fisher Qubit and Agilent Bioanalyzer High Sensitivity DNA Kit (Agilent Technologies Ltd., Didcot, UK). The full protocol is additionally included (S.I. 1).

Sequencing was performed on an Illumina NovaSeq X (Illumina, California, USA) with paired-end 150 reads, to a total read output of 100 Gb per 96 sample pools. All runs included negative no-template controls.

### 2.5 Custom oligonucleotide panel

Libraries were enriched using a novel custom oligonucleotide probe panel, which we designed for this study to cover the full diversity of known human bloodborne viruses (106 species), as well as host markers as positive controls (Fig. 1). The panel additionally contained probes for bacterial and parasite pathogens, which were not examined in this study. The full panel comprised 68,938 double-stranded 120-mer oligonucleotides that targeted the full genome of all viruses <100kb in length, and 20kb of DNA viruses including herpesviruses and poxviruses. The panel was constructed as follows: for each targeted viral species, all available genomes were downloaded from the NCBI Virus database and reduced to clusters representing <10% pairwise genome diversity, using cd-hit-est (Li & Godzik, 2006). Genomes in each cluster were aligned using MAFFT v7 (Katoh et al., 2002), and probes designed against the consensus genomes from each cluster, to constrain probe-target distance above the critical 80% sequence identity limit that maximises enrichment efficiency, following principles outlined in (Bonsall et al., 2015). The panel was designed in-house and synthesised by Twist Bioscience.

**Figure 1.**
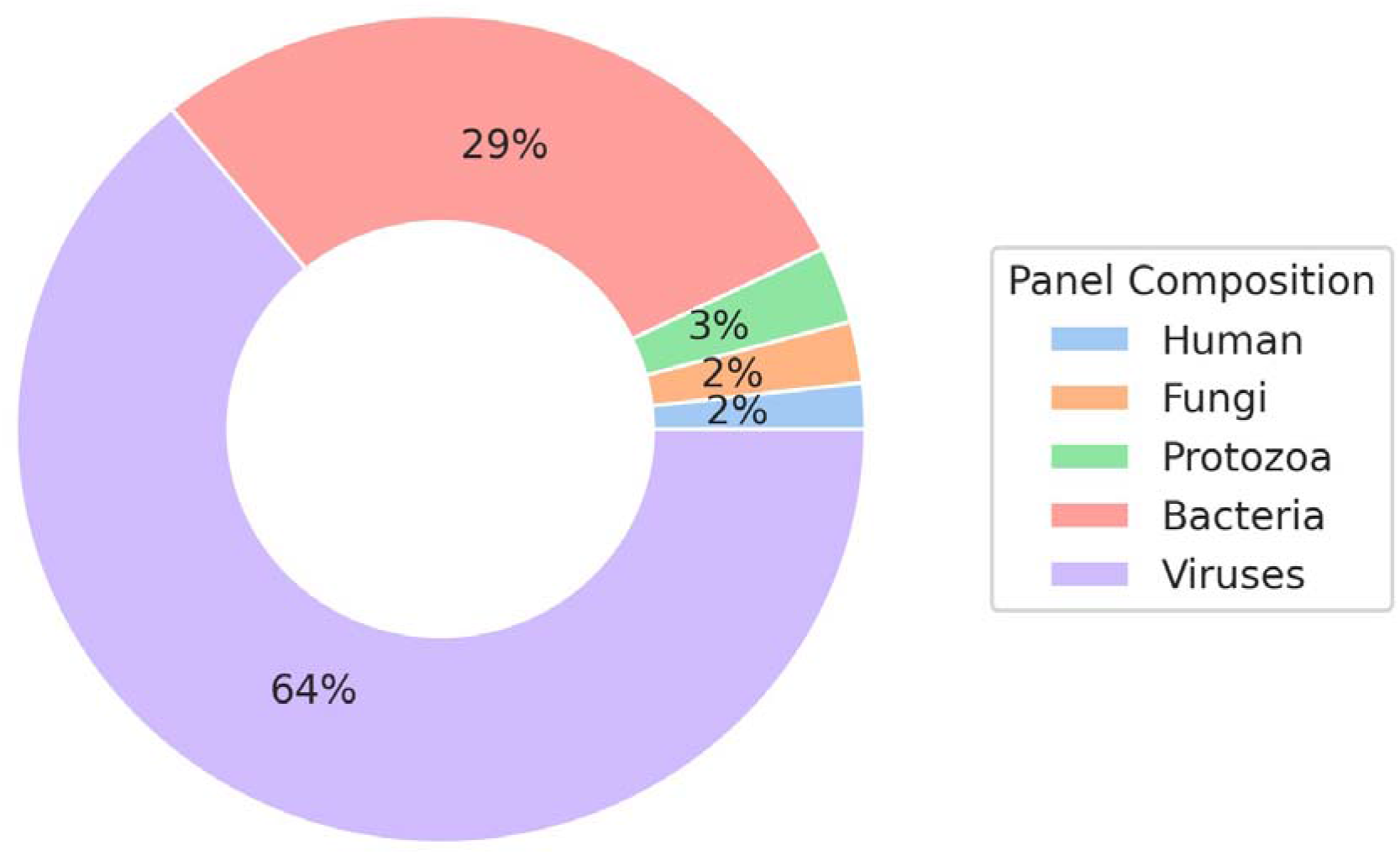
Panel composition by target type, from a total of 68,938 enrichment oligos.

The panel included three host targets: the ABO locus, NADH dehydrogenase-4 and cytochrome C1 oxidase (COX-1), which served as positive internal controls.

### 2.6 Data analysis

#### 2.6.1 Sequencing data processing

NGS data were processed with Castanet (Mayne et al., 2024), a sample- and sequencing-type agnostic pipeline for quality control, read filtering, mapping, read aggregation to specific organisms and consensus sequence deconvolution.

#### 2.6.2 Use of quantification controls

Least squares linear regression was calculated for deduplicated read depth against viral load for quantified controls. As there is a strong linear relationship between these quantities (Bonsall et al., 2020; Lin et al., 2024; Lythgoe et al., 2021; Mayne et al., 2024), coefficient of determination (*R*^2^) values were used to evaluate the relative success of sequencing and estimate viral loads in non-quantified samples.

#### 2.6.3 Limit of detection, sensitivity and specificity

Limits of detection (LOD) were estimated as the lowest detectable viral load above which there were no false negatives; the lowest viral load at which we could be confident in repeatable detection. In cases where LOD could not be determined due to reliable detection at all concentrations, i.e. where no false negatives were reported at any concentration, the LOD was reported as ‘< *x’*, where *x* was the titre of the lowest-concentration sample.

Detection was additionally considered unsuccessful in instances where the virus of interest was found in a sample, but of the genome was recovered.

Sensitivity (Eq. 1) and specificity (Eq. 2) were calculated for the detection of HCV and HEV in clinical samples, where *TP* and *TN* denote true positive and true negatives, and *FP* and *FN* indicate false positive and false negative results. Sensitivity and specificity were used to calculate multi-class receiver operating characteristic (ROC) curves, using the scikit-learn library for Python 3. Three ROC classes were encoded (0: NAT negative, 1: NAT positive, above NGS LOD, 2: NAT positive, below NGS LOD) and macro-average AUCs were calculated by one-versus-one binarization. In other words, all classes were treated equally by evaluating each against all others, recording performance as the average of precision and recall for each set of observations.

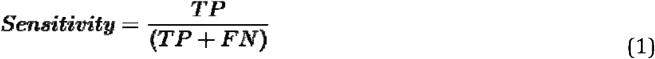

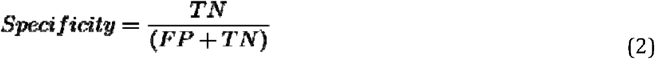

#### 2.6.4 Viral community diversity and prevalence

Diversity of viral communities was calculated as Simpson’s Index, *D* (Eq. 3), where is the number of organisms per unique species and is the total number of organisms.

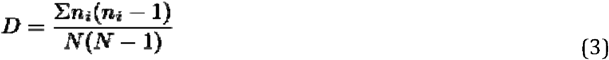

To estimate population prevalence from pooled data, accounting for uncertainty in the number of infections in positive pools, *p*, we used a probabilistic framework, as implemented in EpiTools (Sergeant, E, 2018). Since a pool of 24 plasma donations is only negative when all 24 are negative, we modelled the number of negative pools,, as a binomial random variable with success probability . After adding a uniform prior on, to the observed number of negative pools,, and the number of positive pools,, the posterior distribution of negatives was . We transformed this to the individual-level prevalence,, yielding the full posterior distribution of . Means and credible intervals were obtained from 200,000 samples from *p*. To estimate the distribution of the number of infected individuals per pool, we drew samples from *p*, each time simulating counts from a binomial distribution with 24 trials and success probability *p*.

Where reads for a specific virus were detected in multiple pools, the expected number of individual samples positive for that virus per pool was estimated. The observed frequency per pool, *λ* = −ln (1 − *f*) where *f* was number of observations, was fit to the Poisson distribution for a range of positives per pool (*K*) (Eq. 4).

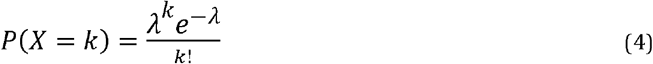

#### 2.6.5 Phylogenetic analysis

Consensus sequences generated by Castanet were aligned using Mafft 7.526 (Katoh et al., 2002), with the L-INS-i algorithm. Maximum likelihood trees were generated using IQ-Tree V2.4.0 (Minh et al., 2020), using the automated ModelFinder selection, with 1000 ultrafast bootstrap replicates.

## 3 Results

### 3.1 NGS method validation

We included control loci in our custom panel design to enable estimates of enrichment level and sequencing quality for each library. This was particularly important as most samples from healthy donors are expected to be negative for the majority of targeted viruses. Inclusion of dilution series of externally quantified viral controls, the NIBSC viral multiple reagent 11/242 (Fig. 2a), allowed us to confirm successful enrichment and estimate relative viral load of clinical samples, based on the number of deduplicated (unique) reads after enrichment (Bonsall et al., 2020; Lin et al., 2024; Mayne et al., 2024). Inclusion of enrichment oligos complementary to host genomic markers, such as the COX-1 gene within the human mitochondrial genome (Fig. 2b), further enabled quantification of amplification above background levels, as the ratio of depth at enriched and unenriched loci. Median amplification exceeded > ×2,000 the background across all human targets in all human-derived samples in this study.

**Figure 2.**
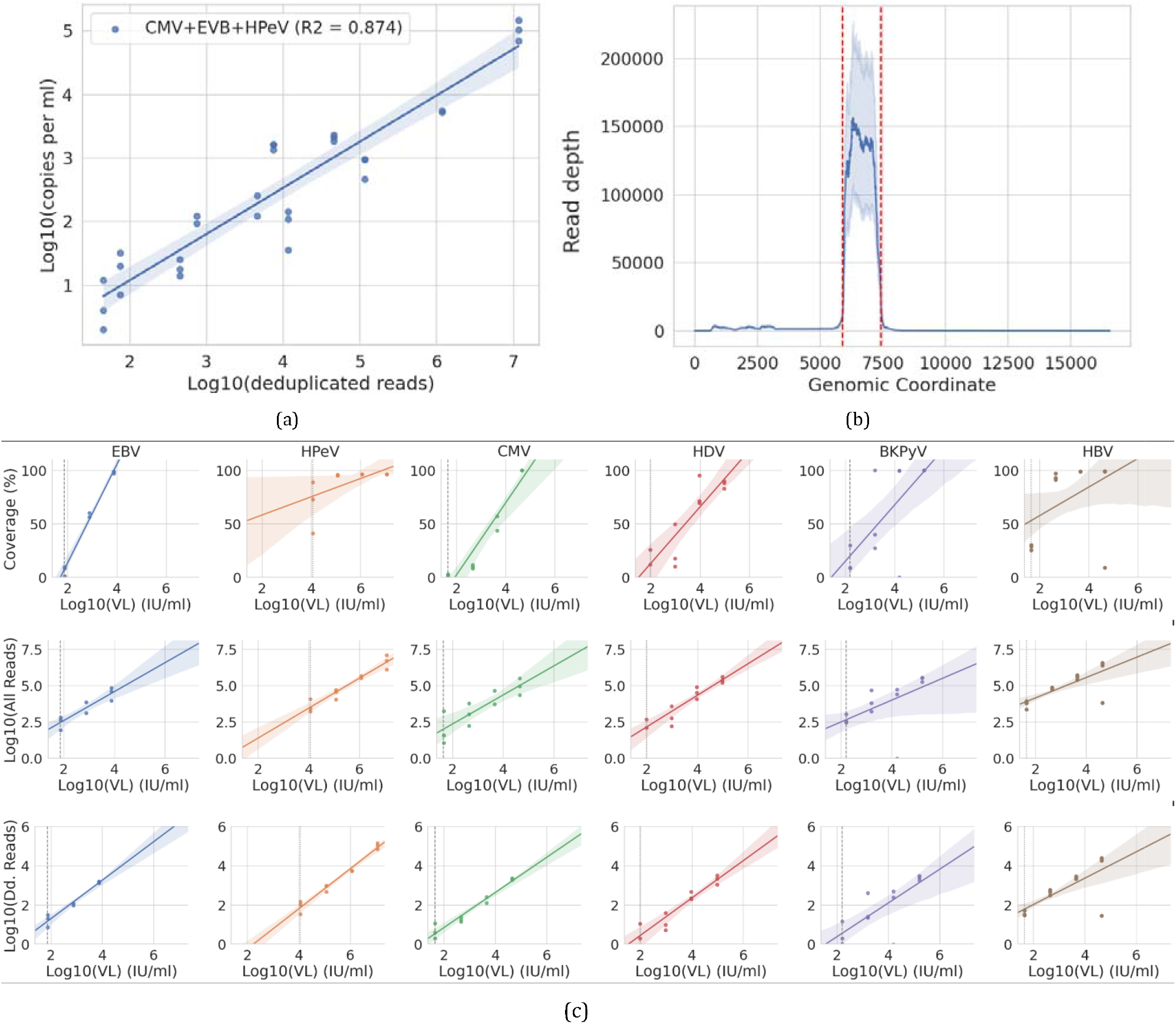
Evaluating capture reactions using quantified and human-derived controls. (a) Regression of viral load against deduplicated read depth (zero values removed) in quantified viral multiplex control demonstrates a strong linear relationship. (b) Read depth is greatly elevated across the enriched region (COX-1 gene; denoted by red lines) of human mitochondrial genome for all pooled plasma samples (showing range, median at midline), which may be used to estimate the degree of amplification across all targets. (c) Coverage, read depth and deduplicated read depth across all quantified controls, with continuous linear regression lines. Vertical lines indicate LODs, where dashed lines are measured limits and dotted indicate less than or equal to.

LODs were calculated as the lowest titre at which sequencing reads were consistently detected, with reference to dilution series for all control samples. Although all viruses within control samples were detected at most dilutions, LODs varied between sample and virus types. LOD values ranged between 43–4235 IU/mL (S.I. 2.2), where HEV was the lowest and HCV was the highest. ROC scores for detection of HCV and HEV in clinical control samples were 0.86 and 0.98, respectively (S.I. 2.2).

### 3.2 Pooled plasma viral content

No significant non-human or non-viral reads were identified in any pooled plasma samples, apart from low-abundance, fragmentary contaminating bacterial and fungal DNA (the ‘kitome’ (Paniagua Voirol et al., 2021; Salter et al., 2014)) that was detectable in all samples including non-template controls, as expected for high-throughput sequencing. A total of 12 separate virus types were found within the 211 pooled plasma samples, most of which were not known to be pathogenic. The majority of pooled samples were found to contain anellovirus and HPgV-1 reads (89% and 72% pools positive, respectively), and reads for a further nine viruses were identified in the minority of samples (Fig. 3). This included herpesviruses HHV6a/b (19/211) and EBV (6), polyomaviruses Merkel cell polyomavirus (MCPyV; 4), BKPyV (1) and human polyomavirus 7 (HPyV7) (1), parvoviruses PARV4 (1) and B19V (1), human gemykibivirus (HuGkV, 5) type 1, HPeV type 3 (1) and HCV type 1a (1). Detection of HPeV was supported by confirmatory PCR.

**Figure 3.**
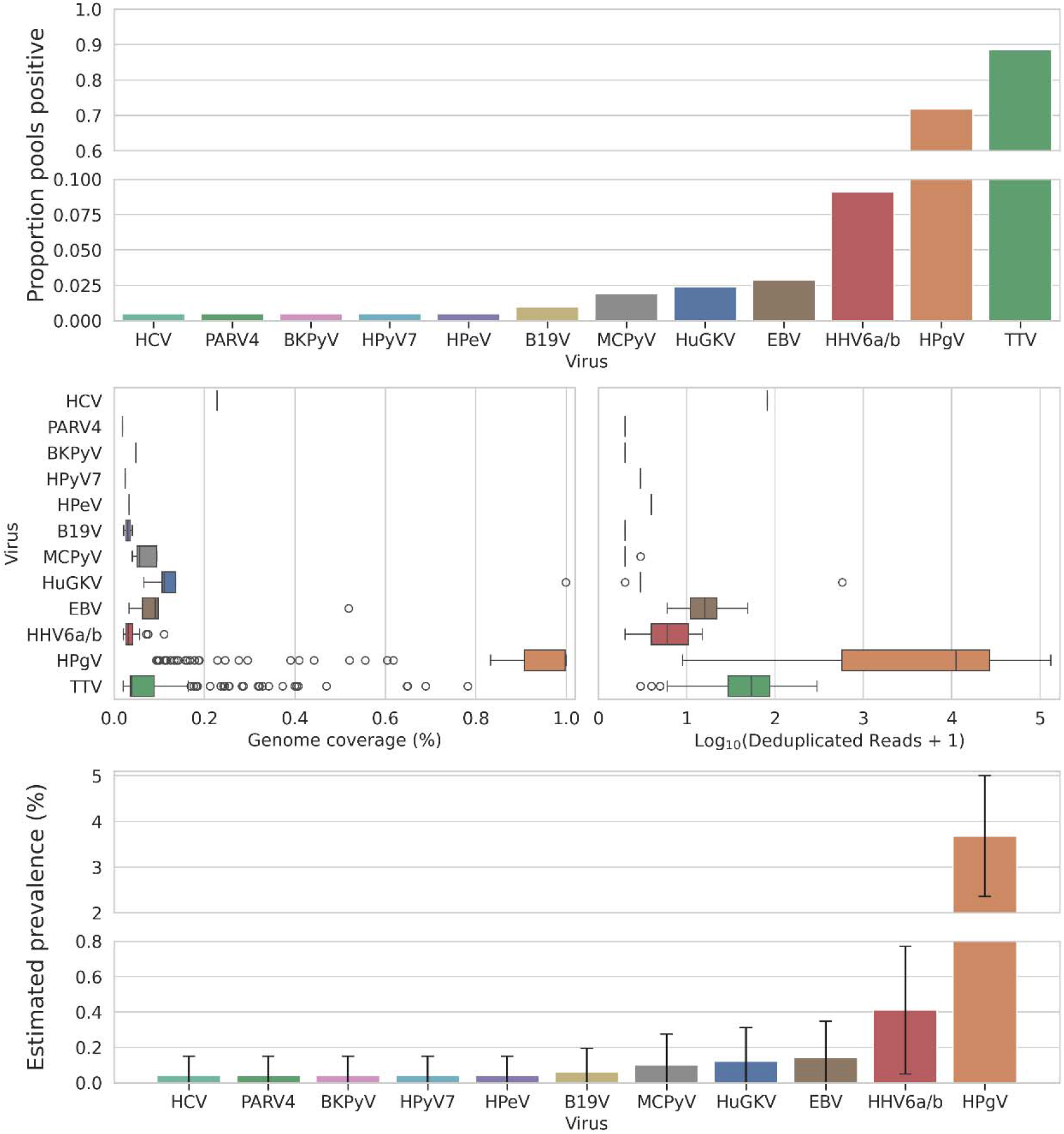
Per virus summaries of prevalence per pool, percentage of genomic positions covered, deduplicated read depth and prevalence per donor inferred from probabilistic framework (with 95% confidence interval) in 211 plasma pools, each comprised from 24 individual donor samples.

The Simpson Diversity Index for the virus community observed was 0.6. Given the near-ubiquity of anelloviruses and HPgV-1 and paucity of other viruses, it was not possible to measure correlation between co-occurrence of specific species.

Each of the 211 plasma pools comprised 150µL of plasma each from 24 individual blood donors, meaning the total number of donors represented was 5,064, approximately 0.63% of the UK donor pool in 2022. Extrapolating incidence rates at the levels of individual donors therefore brings a measure of uncertainty. Statistical modelling indicated virus prevalence estimates range from 0.04%–3.68% (Fig. 3). Modelling was not performed for anelloviruses, which were assumed to be near-ubiquitous (see S. 3.3).

### 3.3 Phylogenetic analyses on non-pathogenic viruses

101 complete HPgV-1 sequences were reconstructed from 101 positive samples. HPgV-1 prevalence was estimated as 3.68% (95% CI 3.02—4.35%) (S.I. 2.3), in line with the expectation that 2–3% of the UK population are viraemic (Rascovan et al., 2016). Consensus sequence reconstruction and haplotype deconvolution using Castanet recovered at most a single complete HPgV-1 genome per pool, though modelling indicated that approximately half of HPgV-1 positive pools would have contained plasma from multiple positive donors (S.I. 2.3). The distribution of genotypes was 97 (97%) genotype 2, 1 (1%) genotype 1, three (3%) genotype 3 (Fig. 4a, S.I. 3), consistent with previous observations that individuals of European origin tend to be infected with genotype 2, whereas genotypes 1 and 3 are more prevalent in Africa and Asia, respectively (Yu et al., 2022).

**Figure 4.**
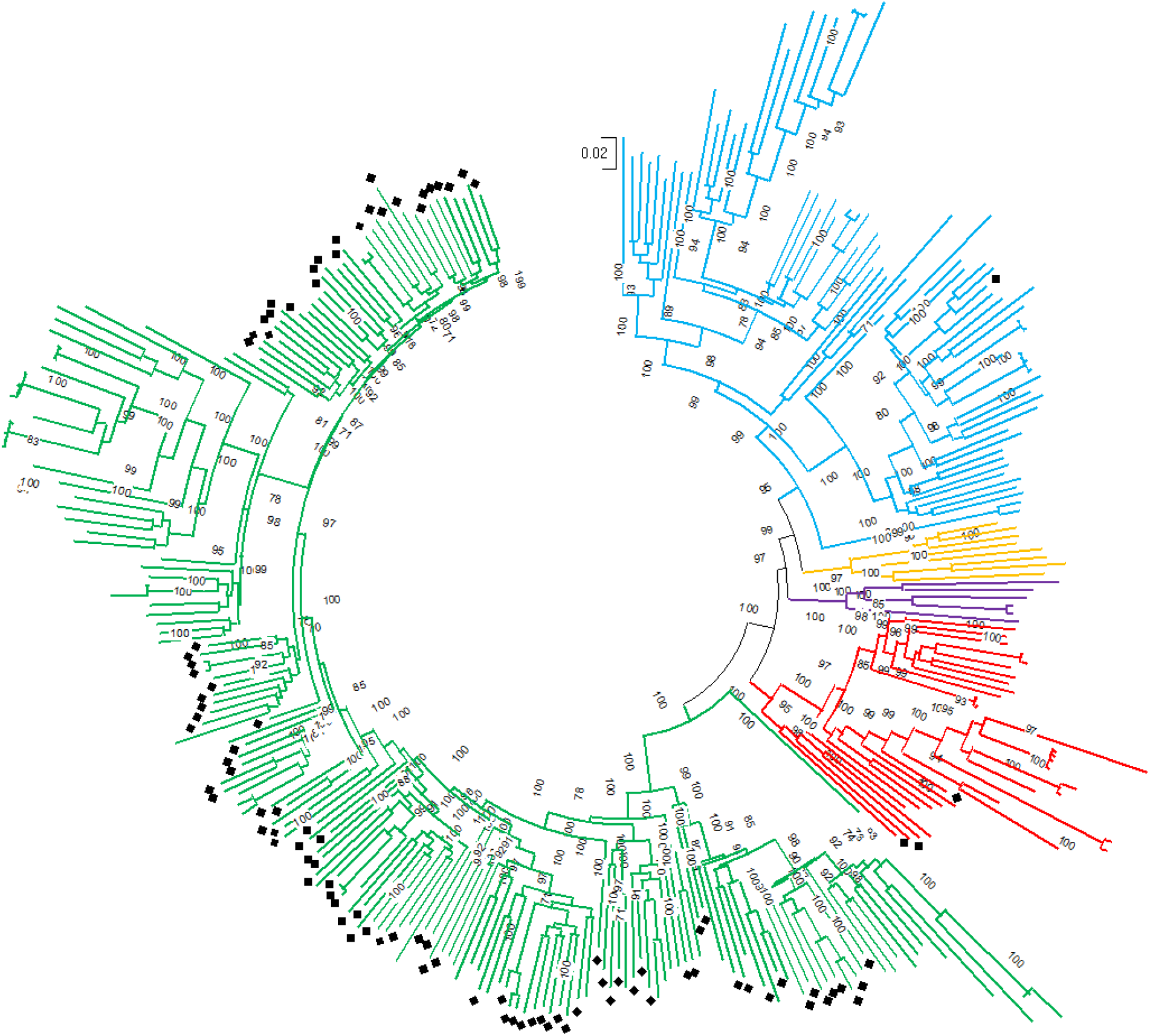
Maximum-likelihood phylogenetic tree for HPgV-1 sequences (*n* = 101, diamonds) reconstructed from donor plasma pools with bootstrap values (values of ≥70% shown) and reference sequences for genotypes 1 (blue), 2 (green), 3 (red), 4 (purple) and 5 (yellow).

No complete anellovirus genomes were recovered, with coverage ranging 2 to 79%. Read totals (log mean of 3019) and genome coverage (log mean 8.5%) across anelloviruses (Fig. 3) were substantially lower than would be expected for the measured viral loads of TTV in 80 representative plasma pools (median 478, inter-quartile range 190-871; S.I. 2.4). The latter values were consistent with the analysis of 1020 individually tested healthy blood donors with a weighted mean of 934 DNA copies/ml calculated from the viral load distributions reported in (Focosi et al., 2020). Quantitative assays for anelloviruses in other anellovirus genera (TTMV, TTMDV) may have led to substantially higher viral loads than determined using the current TTV-specific quantitative assay (Maggi et al., 2003). Genotype assignments for all sequences generated through sequencing (filtered at *>* 10% coverage, *n* = 13) were additionally confirmed through BLASTn searches against the core_nt database. Thirteen partial sequences were recovered, of which nine (⍰70%) were TTV (*Alphatorquevirus*) and four were torque teno midi virus (TTMDV, *Gammatorquevirus*) (S.I. 4). No torque teno mini virus (TTMV, *Betatorquevirus*) sequences were recovered. These results contrast observations made using nested PCR on 40 pools, which revealed all were positive for TTV and TTMV, and 75% (30) were positive for TTMDV.

Only a single complete HuGkV genome was recovered. The remaining positive pools contained partial HuGkV type 1 genome sequences with coverage ranging from 6 to 99%.

## 4 Discussion

This characterisation of plasma from 211 unscreened pools representing over 5,000 donors represents a snapshot of the UK’s healthy plasma virome. The majority of pools screened were unremarkable in our analyses, which should provide some reassurance for blood service professionals and transfusion recipients. While a single pool was found to contain HCV, this would likely have been detected by NHSBT NAT screening and excluded from transfusion; this observation was consistent with the expected donor positivity rate in the UK (∼0.04% here versus 0.03% in (Reynolds et al., 2018)). Large proportions of the population are seropositive for a range of polyomaviruses (Kamminga et al., 2018), parvoviruses (Moosazadeh et al., 2023; Mossong et al., 2008) and parechoviruses (Alam et al., 2025; Westerhuis et al., 2013), but prevalence of viraemia with these viruses in the healthy population is infrequently described in the literature; our findings indicate prevalence estimates for these viruses from 0.04% to 0.10%. Viraemia with pegiviruses (Yu et al., 2022) and anelloviruses (Focosi et al., 2020; Görzer et al., 2023) is expected in healthy individuals at rates of ∼3% and >90% respectively, although the determining factors for an individual being viraemic, either acutely or chronically, are in many cases poorly understood (Moustafa et al., 2017; Rascovan et al., 2016). Our observations add to an underserved area of the literature, as to our knowledge, ours is one of the few large-scale analyses of the plasma virome in the general population (Feng et al., 2023; Moustafa et al., 2017), but the only such recent study in the UK focussing on both DNA and RNA viruses.

Our results are broadly consistent with the findings of a 2017 study (Moustafa et al., 2017) studying the whole blood DNA virome from 8,000 donors in the USA, which found a comparable distribution of polyoma- and herpesviruses, but at higher prevalences than was observed here. These differences may be ascribed to the 2017 study’s use of whole blood, which would increase the proportion of cell-associated material (e.g. herpesviruses) compared with the plasma used here, differences in demographic and sample size, and their inclusion of samples from symptomatic patients. Conversely, we observed an almost identical prevalence of HPgV-1 infection to a recent meta-analysis (Yang et al., 2020) from European cohorts, and the genotype distribution therein was also as expected, i.e. predominantly genotype 2 (Yu et al., 2022). Further, a Chinese study on paired blood-plasma pools from 2023 (Feng et al., 2023) found similar prevalences to the current study (after adjusting for differences in pool size) for anelloviruses, EBV, HHV6, MCPyV and HPgV-1, a lower prevalence of HPeV and higher prevalence of HCV, HCMV, B19V and a variety of other herpes- and papillomaviruses; these differences highlight the variability in virome compositions between geographical regions and blood donor selection policies. The same study found that some viruses were detected in higher quantities in plasma (e.g. herpesviruses) whereas others were less so than in blood, such as anelloviruses.

A full genome sequence and several fragments of a small, widely distributed human DNA virus in the family *Genomoviridae*, genus *Gemybikivirus* (Varsani & Krupovic, 2017) were detected in five plasma pools. HuGKV and members of several other genera such as *Gemycircularvirus* in the family *Genomoviridae* are small DNA viruses with genome sizes ranging from 2-2.4 KB of single stranded, circular DNA encoding capsid and *Rep* genes (Varsani & Krupovic, 2017). Many genera, including gemykibiviruses, show an extraordinarily wide host range, including arthropods, plants, fungi and mammals. Several different divergent genotypes of HuGKV have been characterised in human infections, being reported from blood donors in Brazil (Zucherato et al., 2023), and sporadically in association with varied human clinical disease presentations (Tuladhar et al., 2024; Wang et al., 2019) but without established causation. The five plasma pools in the current study from which HuGKV were detected were all of genotype 1, although this may be an overly conservative estimate of the diversity of plasma-associated HuGKV: the capture probes used for target enrichment were limited to representative genomes of two gemykibivirus genotypes (types 1 and 2) and a single gemycircularvirus. Analogously to limited detection of anelloviruses in pools with viral loads approaching 1000 DNA copies /ml, this limited representation of the very diverse *Genomoviridae* family may have prevented detection of an even wider range of genomoviruses. Nonetheless, the observation of HuGKV at an expected prevalence of >0.12% is notable.

Despite anellovirus reads being present in almost all samples, read depth and coverage were low. Comparison of findings by NGS with consequent quantitative PCR testing revealed no linear relationship between unique reads and TTV viral load, making it impossible to estimate prevalence of anelloviruses using NGS alone. Furthermore, where NGS results indicated minimal representation of TTMV and TTMDV, genotype-specific nested PCR on a subset of 40 pools revealed all were positive for TTV and TTMV, and 75% were positive for TTMDV. Anelloviruses are exceptionally genetically diverse (Arze et al., 2021), with genome sequence diversity not covered fully by representative sequences used to design capture probes used for target enrichment, as is also the case in most comparable studies (Anantharam et al., 2024; Kamitaki et al., 2026; Moustafa et al., 2017). As for genomoviruses, estimation of the true abundance and diversity of anelloviruses may necessitate either much broader target capture enrichment protocols or deeper, shotgun metagenomic sequencing (Arze et al., 2021).

Several limitations are inherent in our study. First, the pooled samples were anonymised, with metadata limited to pooled values for collection year, ethnicity and approximate geographical region of collection, which limits the scope of our analyses to examine individual-level variation. Second, while the sample was large, it still did not permit detection or estimates of the prevalence of rare viruses, such as HAV. Third, although we had extensive controls to demonstrate the efficacy of the NGS method, the use of plasma pools of 24 donations necessarily results in a dilution effect and thus reduces sensitivity for low-titre viruses. However, our use of host targets as positive controls in all samples enabled a robust assessment of overall enrichment, and thus represents a valuable novel approach to designing large hybrid capture panels for screening samples where the majority are expected to be negative for the majority or targeted pathogens. Fourth, no attempt was made to study integration of viral genomes or correlate results with analyses from whole blood.

A slew of recent research has revealed that the blood virome becomes remodelled in various states of health and disease over an individual’s lifetime (Ma et al., 2024; Monaco et al., 2016; Sasa et al., 2025; Thijssen et al., 2020), and in some cases measurements therein may hold predictive value for both diagnosis and prognosis (Görzer et al., 2023). The present study has demonstrated that target enrichment NGS approaches offer a comparatively flexible and cost-effective route to studying the human plasma virome in both health and disease. Future work will consist of comparing larger cohorts from both healthy and non-healthy populations.

## Data Availability Statement

Data are available at European Nucleotide Archives under study accession PRJEB110313.

Analytical scripts and intermediate data are available at https://github.com/MultipathogenGenomics/uk_donor_virome_snapshot_article

## Acknowledgements

KK, RM, KR, SS, BKS, PR, HH, JB, PS and TG acknowledge funding from NIHR, grant number NIHR203338. TG is supported by an Investigator Grant (GNT2025445) from the National Health and Medical Research Council, Australia (NHMRC). KL was supported by the Wellcome Trust and Royal Society (107652/Z/15/Z) and the Li Ka Shing Foundation.

## Disclosures

The authors declare no conflict of interest.

## Supplementary information

1. NGS laboratory protocol
2. NGS method validation additional analyses
  a. Limit of detection, sensitivity, specificity
  b. Virus prevalence estimates
3. Square phylogenetic trees for HPgV-1 and anelloviruses

## Footnotes

1 https://www.nhsbt.nhs.uk/news/nhs-in-england-calls-for-one-million-people-to-donate-blood-to-secure-the-nation-s-bloodsupply/

2 https://www.blood.co.uk/why-give-blood/

## Notes

### Competing Interest Statement

The authors have declared no competing interest.

