## Supplementary Information 2 for "A snapshot of the UK blood donor plasma virome: a retrospective cross-sectional cohort study"

A snapshot of the UK blood donor plasma virome Supplementary Information 2

### 1 Plasma pool demographics

|  | **Female** | **Male** | **Total** |
| --- | --- | --- | --- |
| **Donor type** |  |  |  |
| **Total** | 47.0 | 53.0 | 100.0 |
| **First-time** | 4.1 | 3.5 | 7.6 |
| **Repeat** | 42.9 | 49.5 | 92.4 |
| **Age group** |  |  |  |
| **17-25** | 2.7 | 2.2 | 5.0 |
| **25-34** | 10.1 | 10.4 | 20.5 |
| **35-44** | 9.5 | 10.1 | 19.5 |
| **45-54** | 10.1 | 10.6 | 20.8 |
| **55-64** | 9.5 | 12.6 | 22.0 |
| **65+** | 5.1 | 7.1 | 12.2 |
| **Ethnic group** |  |  |  |
| **Asian/Asian British** | 1.2 | 2.8 | 4.0 |
| **Black/Black British** | 0.9 | 0.8 | 1.7 |
| **Mixed/other** | 1.3 | 1.3 | 2.7 |
| **White** | 43.2 | 47.4 | 90.6 |
| **Unknown** | 0.3 | 0.7 | 1.0 |

Table S2.1.1: Plasma pool donor demographics, as percentages.

### 2 NGS method evaluation

|  | Sensitivity | Specificity | ROC AUC | *R*2 | N detected | LOD |
| --- | --- | --- | --- | --- | --- | --- |
| HCV | 83 | 100 | 0.86 | 0.72 | 10/12 | 4,235 |
| HEV | 100 | 100 | 0.98 | 0.84 | 13/13 | 43 |

Table S2.2.1: Statistics on detection of main blood borne virus in known positive clinical samples. ROC AUC: receiver operating characteristic area under curve; *R*^2^: deduplicated reads against viral load linear regression coefficient of determination; LOD: limit of detection (Log10 IU/ml).

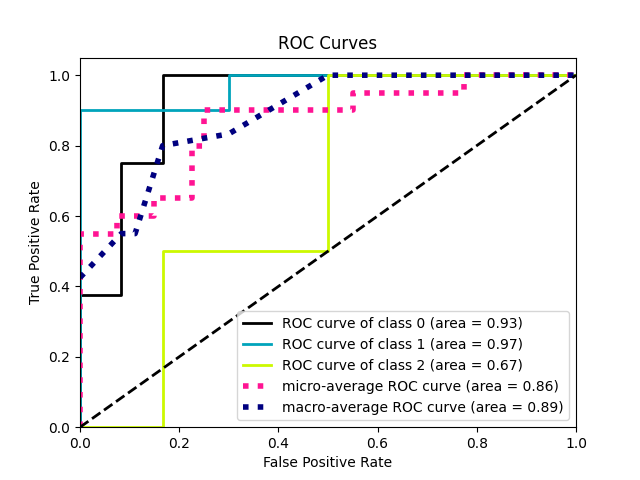

(a) HCV

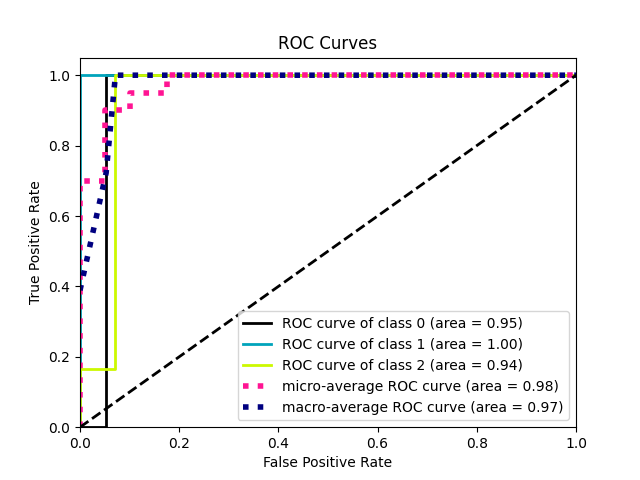

(b) HEV

Fig. S.2.2.1: ROC AUC (one-verses-one macro-average) performance curves for detection of HCV and HEV in quantified clinical samples.

| **Virus** | **Viral load** | $\boldsymbol{R}^{\boldsymbol{2}}$ | **LOD** | **TR D1** | **Pos cov 10** | **DR D1** | **Cov** |
| --- | --- | --- | --- | --- | --- | --- | --- |
| BKV | 5.2 |  |  | 288,924 (85,091) | 5,428 (1) | 2,377 (562) | 1 (0) |
|  | 4.2 | 0.94 | 1,500 | 44,339 (16,650) | 5,408 (26) | 363 (115) | 0.19 (0.11) |
|  | 3.2  2.2 |  |  | 2,991 (2,834)  309 (35) | 1,745 (394)  482 (4) | 20 (6)  2 (1) | 0.3 (0.07)  0.09 (0) |
| HDV | 4.8 |  |  | 260,053 (120,394) | 2,006 (61) | 2,111 (1,004) | 0.94 (0.03) |
|  | 3.8 | 0.9 | *<*60 | 41,619 (35,554) | 1,802 (256) | 284 (140) | 0.79 (0.14) |
|  | 2.8 |  |  | 1,506 (1,992) | 563 (412) | 17 (18) | 0.29 (0.23) |
|  | 1.8 |  |  | 308 (2484) | 395 (152) | 7 (6) | 0.19 (0.10) |
| EBV | 3.9 |  |  | 38,435 (30,319) | 19,322 (134) | 1,526 (143) | 0.98 (0.01) |
|  | 2.9 | 0.99 | 75 | 4,338 (4,283) | 11,153 (635) | 108 (21) | 0.58 (0.03) |
|  | 1.9 |  |  | 378 (267) | 373 (177) | 20 (13) | 0.06 (0.05) |
| CMV | 4.7 |  |  | 139,599 (147,413) | 21,214 (32) | 2,063 (214) | 1 (0) |
|  | 3.7 | 0.99 | 45 | 24,109 (26,513) | 9,019 (3,420) | 190 (97) | 0.51 (0.09) |
|  | 2.7 |  |  | 2,318 (3,003) | 1,758 (857) | 19 (6) | 0.1 (0.02) |
|  | 1.7 |  |  | 621 (1,033) | 158 (199) | 6 (5) | 0.02 (0.01) |
| HPeV | 7.0 |  |  | 6,208,513 (5,569,163) | 7,347 (3) | 107,288 (38,513) | 0.96 (0) |
|  | 6.0 | 0.99 | *<*11,750 | 392,730 (88,225) | 7,340 (8) | 5,397 (168) | 0.96 (0) |
|  | 5.0 |  |  | 33,092 (20,565) | 7,288 (44) | 787 (278) | 0.95 (0.01) |
|  | 4.0 |  |  | 5,567 (5,845) | 4,622 (2,115) | 97 (55) | 0.68 (0.24) |
| HBV | 4.7  3.7  2.7  1.7 | 0.98 | <45 | 2,269,932 (1,113,664)  388,262 (146,591)  64,995 (8,717)  5,458 (3,068) | 3,104 (2)  3,102 (1)  2,906 (61)  885 (75) | 19,407 (7865)  2,569 (415)  442 (134)  39 (14) | 0.99 (0)  0.99 (0)  0.95 (0.02)  0.31 (0.05) |

Table 2.2.2: Statistics on detection of quantified viruses in control samples. Data from dilutions where no reads were identified have been omitted. *R*^2^: deduplicated reads against viral load linear regression coefficient of determination; LOD: limit of detection (copies/ml). TR/DR: total/deduplicated reads; D*x*: mean number of reads at depth *x* (standard deviation); Pos cov: number of genomic positions with coverage at minimum depth 10 (standard deviation); Cov: proportion of genome with coverage at minimum depth 2.

### 3 HPgV-1 prevalence estimates

| **Virus** | **Prevalence** | **95% CI** |
| --- | --- | --- |
| PARV4 | 0.039% | -0.01524, 0.09401 |
| BKPyV | 0.039% | -0.0153, 0.09405 |
| HPyV7 | 0.039% | -0.01549, 0.09429 |
| B19V | 0.059% | -0.00825, 0.1266 |
| HPeV | 0.039% | -0.0153, 0.09405 |
| MCPyV | 0.099% | 0.01178, 0.18649 |
| HuGKV | 0.119% | 0.02345, 0.21533 |
| EBV | 0.139% | 0.03548, 0.24321 |
| HHV6a/b | 0.410% | 0.23059, 0.5912 |
| HPgV-1 | 3.684% | 3.02361, 4.34492 |

Table 2.3.1: Probabilistic estimates with 95% confidence interval for prevalence of each virus observed per donor.
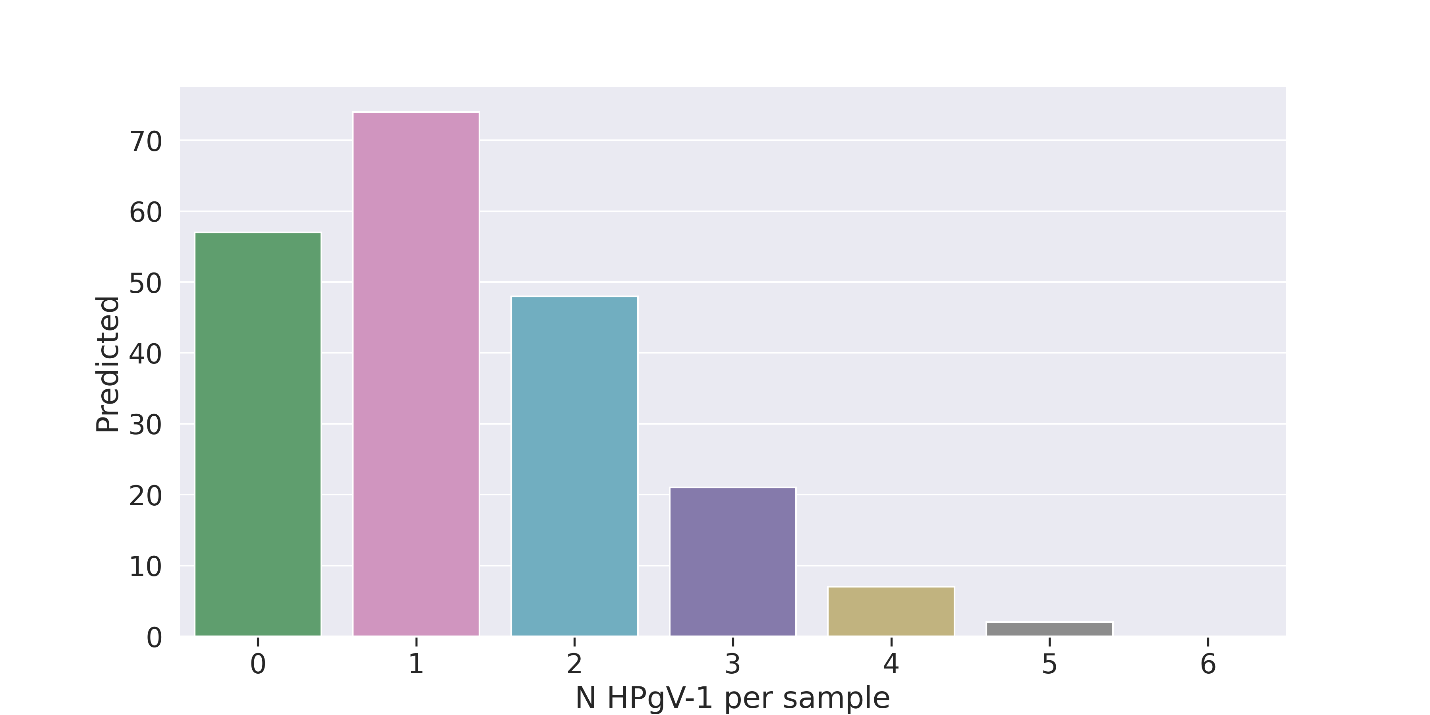
Fig. 2.3.1: Poisson distribution estimates for number of HPgV-1 positive plasma samples per pool.

### 4 TTV PCR

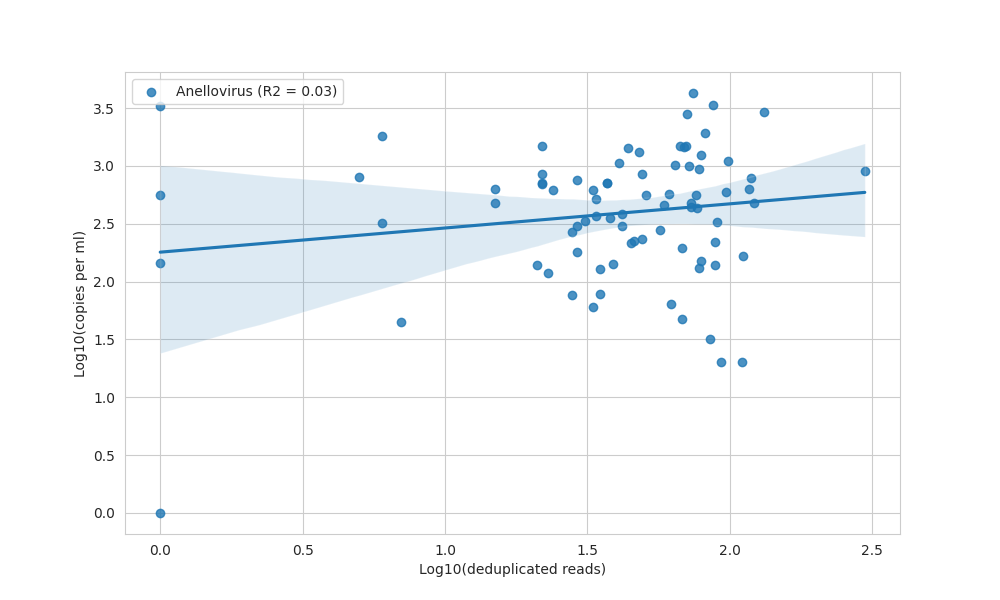

Fig. 2.4.1. Linear regression (OLS) of deduplicated reads against copies per millilitre for anellovirus TTV in 80 pooled plasma samples, showing absence of a linear relationship.

|  | **Log10(Viral Load)** | **Unique Reads** | **Coverage** |
| --- | --- | --- | --- |
| Mean | 2.59 | 55.18 | 8.16 |
| St. Dev. | 0.59 | 42.31 | 10.11 |
| Median | 2.68 | 47.50 | 4.00 |
| IQR Low | 2.28 | 28.00 | 4.00 |
| IQR High | 2.94 | 76.25 | 7.00 |

Table 2.4.1. NGS and viral load descriptive statistics for anelloviruses in 80 pooled plasma samples. Coverage given in percentage of genomic positions.
