## Supplementary figures and images for "A snapshot of the UK blood donor plasma virome: a retrospective cross-sectional cohort study"

### Supplementary Information 3a

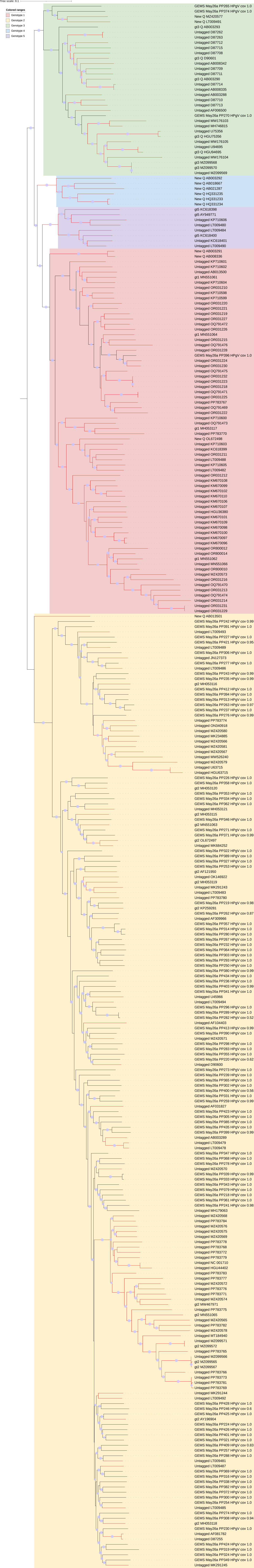

### Supplementary Information 3b

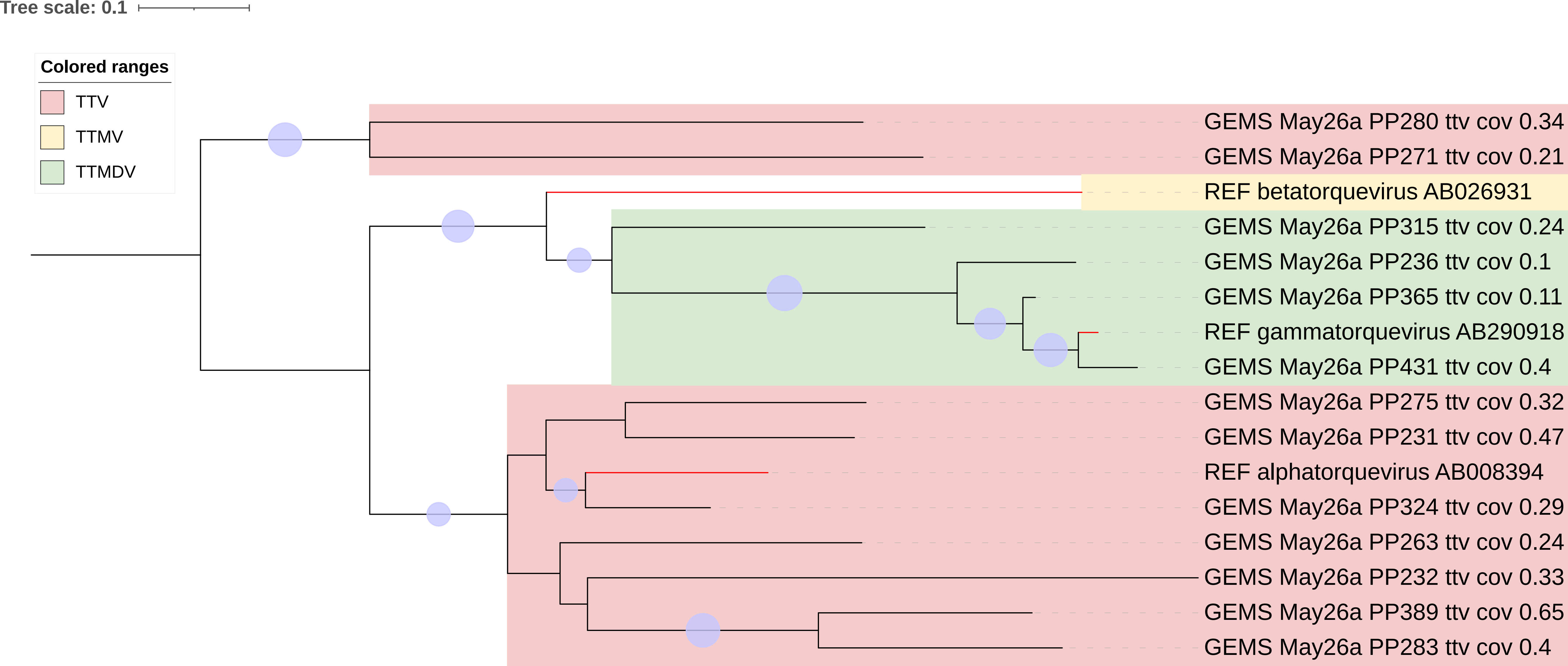
