## Supplementary Information 1 for "A snapshot of the UK blood donor plasma virome: a retrospective cross-sectional cohort study"

GEMS LIBRARY PREP V3

cDNA SYNTHESIS

IMPORTANT: IF USING A PLATE AND PLATE SEALER. SPIN SAMPLES DOWN BETWEEN STEPS TO AVOID CROSS CONTAMINATION ON THE LID

### First strand cDNA synthesis- carry out in PCR set-up hood (~2 hrs)

**Reagents/equipment**

- Superscript IV VILO master mix (200ul)
- Template RNA (8 ul each)
- Nuclease free water (3400 uL)
- Thermocycler
- NEBNEXT Second Strand Synthesis Reaction Buffer (400 ul)
- NEBNext Second Strand Synthesis Enzyme Mix (200 ul)
- First-strand Synthesis Product (10 uL each)
- Ice box
- 96wp cold block
- SPRI/ DNA beads (7200 uL) (take out of fridge to equilibrate whilst 2^nd^ strand synthesis runs)
- Plate Centrifuge
- Magnetic plate rack
- Freshly prepared 80% ethanol (60 ml)

1. Add the following components to an empty RNase-free tube on ice.

| **cDNA first strand reaction** | **1x** | **100x** |
| --- | --- | --- |
| SuperScript IV VILO Master Mix | 2 uL | 200 |
| Template RNA* | 8 uL | - |
| Nuclease-free Water* | 10 uL | 1000 |
| Total | 20 uL | 1200 |

Hint: aliquot 150 ul into each well of a PCR strip to pipette using a multichannel

2. Add 12 uL to each well of a 96wp on an ice block
3. Add 8uL extracted RNA from each sample to each well
4. Gently mix and incubate in thermocycler at:

- 25°C for 10 minutes
- 50°C for 10 minutes
- 85°C for 5 minutes.

Use cDNA immediately for Second Strand Synthesis or store at –20°C for up to one week, or –70°C for long term storage.

### Second strand cDNA synthesis (~4 hrs)

1. Assemble the second strand cDNA mastermix on ice

| **cDNA Mastermix** | **1x** | **100x** |
| --- | --- | --- |
| Nuclease-free Water | 24 uL | 2400 |
| NEBNext Second Strand Synthesis Reaction Buffer | 4 uL | 400 |
| NEBNext Second Strand Synthesis Enzyme Mix | 2 uL | 200 |
| First-strand Synthesis Product | 10 uL | - |
| Total | 40 uL | 3000 |

Hint: pipette out master mix into PCR strip to use multichannel

2. Add 30 uL to each well of a 96wp on ice
3. Add 10 uL of first strand product to each well, mix thoroughly by pipetting the reaction up and down at least 10 times.
4. Incubate in a thermocycler for 1 hour at 16°C with the heated lid set at ≤ 40°C (or off). Meanwhile get DNA beads out to equilibrate
5. Vortex SPRIselect Beads or NEBNext Sample Purification Beads to resuspend
6. Add 72 μl (1.8X) of resuspended beads to the second strand synthesis reaction (~40 μl). Mix well on a vortex mixer or by pipetting up and down at least 10 times.
7. Incubate for 5 minutes at room temperature.
8. Briefly plate the tube in a centrifuge to collect any sample from the sides of the tube.
9. Place the tube on a magnetic rack to separate beads from the supernatant.
10. After the solution is clear, carefully remove and discard the supernatant. Be careful not to disturb the beads, which contain DNA. (Caution: Do not discard beads)
11. Add 200 μl of freshly prepared 80% ethanol to the tube while in the magnetic stand. Incubate at room temperature for 30 seconds, and then carefully remove and discard the supernatant
12. Repeat Step 2.5 once for a total of 2 washing steps.
13. Air dry the beads for up to 5 minutes while the tube is on the magnetic rack with the lid open.

(Caution: Do not overdry the beads. This may result in lower recovery of DNA target. Elute the samples when the beads are still dark brown and glossy looking, but when all visible liquid has evaporated. When the beads turn lighter brown and start to crack they are too dry.)

14. Remove the tube from the magnet. Elute the DNA target from the beads by adding 11 μl 0.1XTE Buffer (or water) to the beads. Mix well on a vortex mixer or by pipetting up and down ten times. Briefly spin the tube and incubate for 2 minutes at room temperature. Place the tube on the magnetic rack until the solution is clear.
15. Remove 5 μl of the supernatant and transfer to a clean nuclease-free PCR plate. Remove another 5 ul and place in another PCR plate. Wash delicately: take care to not dry out beads, especially when doing large quantity of samples. Note: If you need to stop at this point in the protocol, samples can be stored at –20°C.
16. Use the Qubit dsDNA high sensitivity Quantitation Assay to determine the concentration of your dscDNA/gDNA samples. (Although we record the concentration of our DNA we DO NOT adjust the our input volumes as total DNA concentration is not indicative of viral/microbial DNA concentration. Qubit may detect little/no cDNA as concentration will be very low).

(Qubit: GUT control, stem cells, pooled plasma and NXC)

LIBRARY PREP

### Day 1:

### 1. DNA FRAGMENTATION, END REPAIR, AND dA-TAILING (1hr)

**Reagents/equipment**

- cDNA
- Molecular biology grade water (chilled)
- From the Twist Library Preparation EF Kit 1:

• 5x Fragmentation Enzyme

• 10x Fragmentation Buffer

- PCR plate
- Thermocyler
- Ice box
- 96wp cold block

**Protocol**

- Thaw 5x Fragmentation Enzyme and gDNA samples on ice, then mix by flicking the tube with a finger.
- Thaw 10x Fragmentation Buffer on ice, then mix by pulse vortexing for 2 seconds.
- Do serial dilution of cDNA samples that need it
- Program the thermal cycler with the following conditions:

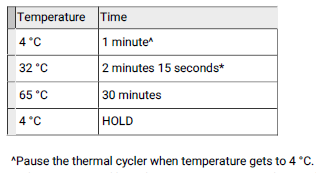

Hint: thermocycler program saved as ‘FRAGMENTATION’ in left most thermocycler

- Set the temperature of the heated lid to 105°C . Start the program to pre-chill the thermal cycler (4 °C). Pause when it gets to 4 °C.
- Add **5 μl (OR, 10 μl IF DOING DOUBLE TEMPLATE**) of each diluted cDNA sample into a well of a 96-well thermal cycling plate on the cold block.
- Prepare an enzymatic fragmentation master mix in a tube on ice. Use the volumes listed below. Mix thoroughly by gentle pipetting. (**REMEMBER TO INCREASE QUANTITY IF DOING DOUBLE TEMPLATE**)

| **Enzymatic frag master mix** | **1x** | **100x** |
| --- | --- | --- |
| Water (chilled) | 15 μl |  |
| 10x Fragmentation buffer | 2.0ul |  |
| 5x Fragmentation Enzyme | 3.0ul |  |
| Total | 20 μl |  |

- Add **20 μl** **(OR, 40 μl IF DOING DOUBLE TEMPLATE**) enzymatic fragmentation master mix to each 5 μl cDNA sample well or tube and mix well by gentle pipetting. Cap the tube and keep the reaction on ice.
- Pulse-spin the sample plate or tubes and immediately transfer to the pre-chilled thermal cycler.
- When the thermal cycler program is complete and the sample block has returned to 4 °C, remove the samples from the block and place on ice.

NOTE: While the thermal cycler program is running, prepare the reagents for next step

### 2. LIGATE TWIST UNIVERSAL ADAPTERS AND PURIFY (1hr)

**Reagents/equipment**

- Thaw on ice: From the Twist Library EF Kit 1:
- Twist **Universal Adapters (TUBE NOT PLATE)**
- DNA Ligation Master Mix
- DNA purification beads (4000 ul)
- Prepare 1 ml 80% ethanol for each sample
- Equilibrate DNA Purification Beads to room temperature for at least 30 minutes
- Program a thermal cycler to incubate the samples at 20 °C with the heated lid set to minimum temperature or turned off. Start the program so that the cycler is at 20 °C when the samples are prepared.

**Protocol**

- Ensure all prep has been done
- Add **2.5 μl** **(OR, 5.0 μl IF DOING DOUBLE TEMPLATE**)Twist Universal Adapters into each sample well containing the dA-tailed DNA fragments
- Mix gently by pipetting 10x and keep on ice (do not vortex)
- Prepare the ligation master mix (below) on ice. (**REMEMBER TO INCREASE QUANTITY IF DOING DOUBLE TEMPLATE**)

| **LIGATION MASTER MIX** | **N=1** | **N=100** |
| --- | --- | --- |
| DNA ligation mix | 10ul | 100 |
| Total | 10 ul | 1125 |

- Add **10** **μl** **(OR, 20 μl IF DOING DOUBLE TEMPLATE**) of the master mix to the sample and mix by gentle pipetting 10x
- Incubate the ligation reaction at 20 °C for 15 minutes in the thermal cycler, then move the samples to the bench top. Proceed to the Purify step.

NOTE: While the thermal cycler program is running, prepare the reagents for the next section (PCR Amplify Using TWIST UDI Primers, Purify, and Perform QC – part 3)

- Vortex the pre-equilibrated DNA Purification Beads until well mixed.
- Add 30 μl of homogenized DNA Purification Beads to each ligation sample. Mix well by vortexing.
- Incubate the samples for 5 minutes at room temperature
- Place the samples on a magnetic plate for 1 minute or until the supernatant is clear
- The DNA Purification Beads form a pellet, leaving a clear supernatant. Without removing plate or tubes from the magnetic plate, remove and discard the supernatant.
- Wash the bead pellet by gently adding 150 μl freshly prepared 80% ethanol (do not disturb the pellet). Incubate for 1 minute, then remove and discard the ethanol.
- Repeat the wash once, for a total of two washes. Keeping the plate on the magnet the whole time
- Carefully remove all remaining ethanol with a 10-μl pipet, making sure not to disturb the bead pellet. NOTE: Before pipetting, the bead pellet may be briefly spun to collect ethanol at the bottom of the plate or tube and returned to the magnetic plate.
- Air-dry the bead pellet on the magnetic plate for 5 minutes or until the bead pellet is dry. Do not overdry the bead pellet
- Remove the plate or tubes from the magnetic plate and add 8.5 μl water / Buffer EB to each sample. Mix by pipetting until homogenized
- Incubate at room temperature for 2 minutes
- Place the plate or tubes on a magnetic plate and let stand for 3 minutes or until the beads form a pellet
- Transfer 7.5 μl of the supernatant containing the ligated and indexed libraries to a clean thin walled PCR 0.2-ml strip-tube or well of a 96-well thermal cycling plate, making sure not to disturb the bead pellet

Keep bead plate in -20 in case we need to go back to. Label ligated adapters.

### 3. PCR Amplify Using TWIST UDI Primers, Purify, and Perform QC (1.5hrs)

Amplify the adapted gDNA libraries with Twist UDI Primers, purify them, and perform quality

control (QC) analysis

**Reagents/equipment**

- Ligated adapted libraries from previous step
- 80% ethanol
- Equilibrated DNA purification beads (2500 ul)
- Molecular grade water
- Equinox library Amp mix (2x)
- Twist UDI primers (from the twist universal adapter system)

IMPORTANT: Use of Amplification Primers, ILMN tubes 100220, 100583 contained in the Twist Library Preparation EF Kit 1 are **not** required. Using these primers with the Twist Universal Adapter System will result in a failed PCR amplification.

**Protocol**

- Thaw **Twist UDI primers** (**plate** with single use primers) and Equinox library Amp mix (2x) on ice
- Prepare the thermocycler

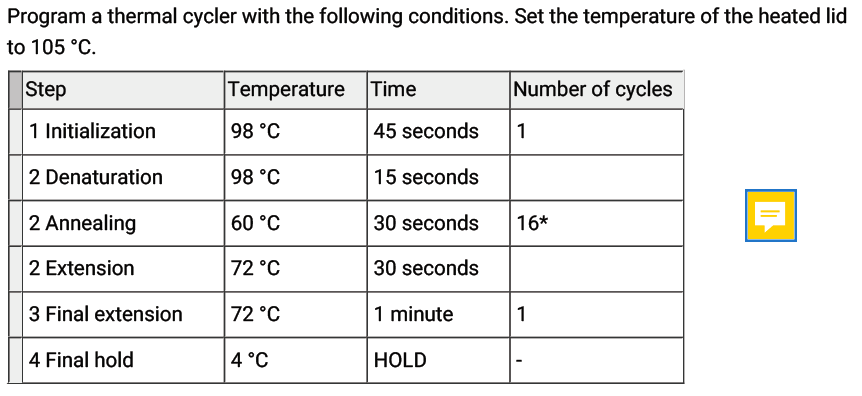

‘SG UDI’ program saved

- Add 5 μl of Twist UDI Primer from the provided 96-well plate to each of the gDNA libraries and mix well by gentle pipetting
- Add 12.5 μl of Equinox Library Amp Mix (2x) to the gDNA libraries and mix well by gentle pipetting.
- Pulse-spin sample plate or tube and immediately transfer to the thermal cycler. Start the program
- Remove the sample(s) from the block when the thermal cycler program is complete. Proceed to purification
- Vortex the pre-equilibrated DNA Purification Beads until mixed.
- Add 25 μl (1x) of homogenized DNA Purification Beads to each ligation sample from Step 3.5. Mix well by vortexing.
- Incubate the samples for 5 minutes at room temperature.
- Place the samples on a magnetic plate for 1 minute
- The DNA Purification Beads form a pellet, leaving a clear supernatant. Without removing plate or tubes from the magnetic plate, remove and discard the supernatant
- Wash the bead pellet by gently adding 150 μl freshly prepared 80% ethanol (do not disturb the pellet), incubate for 1 minute, then remove and discard the ethanol
- Repeat this wash once, for a total of two washes, while keeping the samples on the magnetic plate.
- Carefully remove all remaining ethanol with a 10-μl pipet, making sure not to disturb the bead pellet.
- Air-dry the bead pellet on the magnetic plate for 5 minutes or until the bead pellet is dry. Do not overdry the bead pellet
- Remove the plate or tubes from the magnetic plate and add 11 μl water, 10 mM Tris-HCl pH 8, or Buffer EB to each sample. Mix by pipetting until homogenized
- Incubate at room temperature for 2 minutes.
- Place the plate or tubes on a magnetic plate and let sta10/11nd for 3 minutes or until the beads form a pellet
- Transfer 10 μl of the clear supernatant containing the Amplified Indexed Libraries to a clean thin-walled PCR 0.2-ml strip-tube or well of a 96-well thermal cycling plate, making sure not to disturb the bead pellet
- OPTIONAL: Quantify and validate the size range of each library using the Thermo Fisher Scientific Qubit dsDNA Broad Range Quantitation Assay (2ul)

**STOPPING POINT**: If not proceeding immediately to a Twist Target Enrichment System, store the amplified indexed libraries at –20°C. Seal plate very well!

### 4. PREPARE LIBRARIES FOR HYBRIDIZATION (2 hrs)

**Reagents/equipment**

- Speedy vac
- Libraries

**Protocol**

- Transfer 3 ul from each amplified indexed library to an indexed library pool reaction tube
- Pulse-spin the indexed library pool tube(s) to minimize the amount of bubbles present.
- Dry the indexed library pool(s) using a vacuum concentrator using low or no heat. Depending on volume, expect to take 90-120 min. Check regularly to ensure samples aren’t over-dried. N.b. speedy vac can be force opened by inserting something thin into the lock release, but make sure vacuum has largely dropped first.

STOPPING POINT: If not proceeding immediately to step 2, store the dried indexed library pool at -20 °C for up to 24 hours.

### 5. HYBRIDIZE CAPTURE PROBES WITH POOLS (1hr + 16hrs overnight)

IMPORTANT: Use 96 well plate (tubes will melt)

**Reagents/equipment**

- Indexed library pool
- Twist custom panel
- Twist custom secondary (spike-in) panel(s) (optional)

From Twist Hybridization Reagents:

- Hybridization Mix
- Hybridization Enhancer

From Twist Universal Blockers:

- Universal Blockers
- Blocker Solution (If using a non-human capture panel, replace with species-specific

blocking solution, not provided)

**Protocol**

- Ensure prep is done: thaw all required reagents on ice, then pulse-vortex for 2 seconds to mix and then pulse-spin.
- Set a heat block to 65 °C.
- Program a 96-well thermal cycler to 95 °C and set the heated lid to 105 °C.
- Heat the Hybridization Mix at 65 °C in the heat block for 10 minutes, or until all precipitate is dissolved, then cool to room temperature on the benchtop for 5 minutes
- Prepare a **probe solution** in a clean thin-walled PCR 0.2-ml strip-tube as indicated in the table below. Mix by flicking the tube(s).

| **Probe solution** | **Volume** |
| --- | --- |
| Hybridization mix | 10 ul |
| Twist/custom panel (oligos) | 2 ul |
| Water OR optional spike-in | 0—2 ul |
| Total | 14 ul |

Notes: Hybridization Mix is very viscous. Pipette slowly to ensure accurate pipetting.

Small white particles may be present in the Twist Fixed or Custom Panel tube(s). This will

not affect the final capture product.

- Resuspend the dried indexed library pool by adding the reagents described below. Mix by flicking the tube(s).

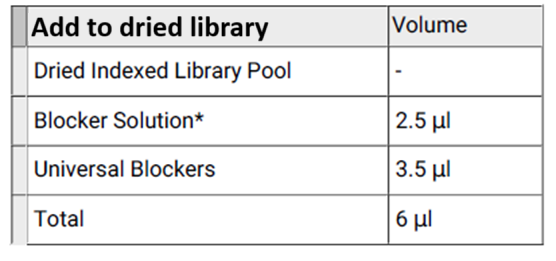

- Heat the **probe solution** to 95 °C for 2 minutes in a thermal cycler with the lid at 105 °C, then immediately cool on ice for 5 minutes
- While probe solution is cooling on ice, heat the tube containing the resuspended indexed library pool at 95 °C for 5 minutes in a thermal cycler with the lid at 105 °C, then equilibrate both the probe solution and resuspended indexed library pool to room temperature on the benchtop for 5 minutes
- Vortex and spin down the probe solution, then transfer the entire volume of the probe solution to the resuspended indexed library pool. Mix well by vortexing
- Pulse-spin the tube(s) to ensure all solution is at the bottom of the tube(s).
- Add 15 μl Hybridization Enhancer to the top of the entire capture reaction
- Pulse-spin the tube(s) to ensure there are no bubbles present. IMPORTANT: Seal the tube(s) tightly to prevent excess evaporation over the 16-hour incubation
- Incubate the hybridization reaction at 70 °C for 16 hours in a thermal cycler with the lid at 85°C. NOTE: Halting hybridization between 15–17 hours will not affect downstream capture quality.

### Day 2:

**N.b. do not conduct any work with capture libraries in the PCR room**

### 1. BIND HYBRIDIZED TARGETS TO STREPTAVIDIN BEADS (1.5-2 hrs)

**Plan for 30 minutes of preparation before removal from overnight thermocycler step**

**Reagents/equipment**

- Hybridization reaction from before

From the Twist Hybridization Reagents:

- Amplification Primers

From the Twist Wash Buffers:

- Binding Buffer
- Wash Buffer 1
- Wash Buffer 2

From Twist Binding and Purification Beads or Twist Dry Down Beads:

- Streptavidin Binding Beads
- DNA Purification Beads

**Protocol**

Preheat the following tubes at 48°C until any precipitate is dissolved:

- Binding Buffer
- Wash Buffer 1
- Wash Buffer 2

For each hybridization reaction:

- Equilibrate 400 μl Binding Buffer to room temperature
- Equilibrate 100 μl Wash Buffer 1 to room temperature
- Leave 350 μl Wash Buffer 2 at 48°C

Thaw on ice (these are used LAST so can be thawed near the end):

- •Equinox Library Amp Mix (2x)
- •Amplification Primers
- Equilibrate DNA Purification Beads (from the Twist Binding and Purification Beads or Twist Dry Down Beads) to room temperature for at least 30 minutes
- Equilibrate the Streptavidin Binding Beads to room temperature for at least 30 minutes
- Make sure that if you are planning to transfer your reagents into a single tube prior to step 10, ensure your strip has 2 tubes in it, which will prevent it from spinning when on the magnetic plate.
- Vortex the pre-equilibrated Streptavidin Binding Beads until mixed
- Add 50 μl Streptavidin Binding Beads to a 1.5-ml microcentrifuge tube.
- Add 100 μl Binding Buffer and mix by pipetting.
- Place the plate on a magnetic stand for 1 minute, then remove and discard the clear
- supernatant. Make sure to not disturb the bead pellet. Remove the tube from the magnetic stand.
- Repeat the wash with 100 ul binding buffer two more times for a total of three washes. (resuspend beads in binding buffer each time)
- After removing the clear supernatant from the third wash, add a final 100 μl Binding Buffer and resuspend the beads by vortexing until homogenized.
- After the 16 hour hybridization is complete, open the thermal cycler lid and directly transfer the volume of each hybridization reaction into a corresponding tube of washed Streptavidin Binding Beads from. Mix by pipetting and flicking.

**IMPORTANT:** Rapid transfer directly from the thermal cycler at 70°C is a critical step for minimizing off-target binding. Do not remove the tube(s) of hybridization reaction from the thermal cycler or otherwise allow it to cool to less than 70°C before transferring the solution to the washed Streptavidin Binding Beads. Allowing to cool to room temperature for less than 5 minutes will result in as much as 10–20% increase in off-target binding.

- Mix the tube(s) of the hybridization reaction with the Streptavidin Binding Beads for 30 minutes at room temperature on a shaker, rocker, or rotator at a speed sufficient to keep the solution mixed. **Do not vortex. (350 rpm)**
- Remove the tube(s) containing the hybridization reaction with Streptavidin Binding Beads from the mixer and pulse-spin to ensure all solution is at the bottom of the tube(s).
- Place the tube(s) on a magnetic stand for 1 minute. Remove and discard the clear supernatant including the Hybridization Enhancer. Do not disturb the bead pellet.
- Remove the tube(s) from the magnetic stand and add 100 μl Wash Buffer 1. Mix by pipetting.
- Pulse-spin to ensure all solution is at the bottom of the tube(s).
- Transfer the entire volume from (~100 μl) into a new 96 well plate (NOT 1.5ml tubes as the thermocycler will melt them at this temperature), one per hybridization reaction. This step is important for removing non-specific binding.
- Place the tube(s) on a magnetic stand for 1 minute.
- Remove and discard the clear supernatant. Make sure to not disturb the bead pellet.
- Remove the tube(s) from the magnetic stand and add 100 μl of 48°C Wash Buffer 2. Mix by pipetting, then pulse-spin to ensure all solution is at the bottom of the tube(s).
- Incubate the tube(s) for 5 minutes at 48°C.
- Place the tube(s) on a magnetic stand for 1 minute.
- Remove and discard the clear supernatant. Make sure to not disturb the bead pellet.
- Repeat the wash two more times for a total of three washes. (repeat 17-20, resuspend the beads)
  - Wash 2
  - Wash 3
- After the final wash, use a 10 μl pipette to remove all traces of supernatant. Proceed immediately to the next step. Do not allow the beads to dry. **NOTE**: Before removing supernatant, the bead pellet may be briefly spun to collect supernatant at the bottom of the tube and returned to the magnetic plate.
- Remove the tube(s) from the magnetic stand and add 22.5 μl water. Mix by pipetting until homogenized, then incubate this solution, hereafter referred to as the Streptavidin Binding Bead slurry, on ice.

### 2. POST-CAPTURE PCR AMPLIFY, PURIFY, AND PERFORM QC (1.5 -2 hrs)

**Reagents/equipment**

- Streptavidin Binding Bead slurry (from before)
- Ethanol
- Molecular biology grade water

Reagents thawed and equilibrated:

- Equinox Library Amp Mix (2x)
- Amplification Primers
- DNA purification beads
- Agilent Bioanalyzer High Sensitivity DNA Kit (or equivalent)
- Thermo Fisher Scientific Qubit dsDNA High Sensitivity Quantitation Assay.
- SPRISelect beads

**Protocol**

- Prepare 1800 μl fresh 80% ethanol for each sample to be processed
- Program a thermal cycler with the following conditions. Set the heated lid to 105°C
- If the Streptavidin Binding Bead slurry has settled, mix by pipetting
- Transfer 11.25 μl of the Streptavidin Binding Bead slurry to a 0.2-ml thin-walled PCR strip-tube(s) or plate. Keep on ice until ready to use in the next step. Store the remaining water/Streptavidin Binding Bead slurry at –20°C for future use.
- Prepare a **PCR mixture** by adding the following reagents to the tube(s) containing the Streptavidin Binding Bead slurry. Mix by pipetting.

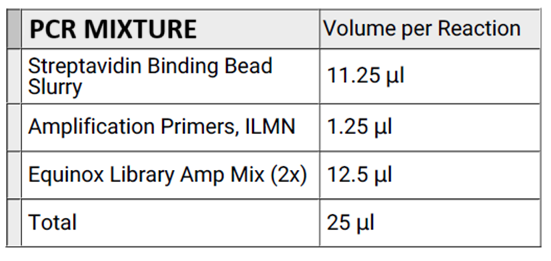

- Pulse-spin the tubes, transfer them to the thermal cycler and start the cycling program.

| **STEP** | **TEMP** | **TIME** | **CYCLES** |
| --- | --- | --- | --- |
| Initialization | 98°C | 45 seconds | 1 |
| Denaturation | 98°C | 15 seconds |  |
| Annealing | 60°C | 30 seconds | 9 |
| Extension | 72°C | 30 seconds |  |
| Final Extension | 72°C | 1 minute | 1 |
| Final Hold | 4°C | HOLD | 1 |

- When the thermal cycler program is complete, remove the tube(s) from the block and return to ice.
- Transfer from 96 well plate back to tube, again ensuring your strip has at least 2 tubes in it to prevent them from spinning in the magnetic plate. Vortex the pre-equilibrated DNA Purification Beads until well mixed
- Add 25 μl (1.0x) homogenized DNA Purification Beads to the tube. Mix well
- by vortexing. NOTE: It is not necessary to recover supernatant or remove Streptavidin Binding Beads from the amplified PCR product.
- Incubate for 5 minutes at room temperature.
- Place the tube(s) on a magnetic plate for 1 minute or until the supernant is clear.
- The DNA Purification Beads form a pellet, leaving a clear supernatant. Without removing the plate or tube(s) from the magnetic plate, remove and discard the clear supernatant
- Wash the bead pellet by gently adding 150 μl freshly prepared 80% ethanol (do not disturb the pellet). Incubate for 1 minute, then remove and discard the ethanol
- Repeat this wash once, for a total of two washes, while keeping the tube on the magnetic plate.
- Carefully remove all remaining ethanol using a 10 μl pipette, making sure to not disturb the bead pellet. NOTE: Before pipetting, the bead pellet may be briefly spun to collect ethanol at the bottom of the plate or tube and returned to the magnetic plate.
- Air-dry the bead pellet on the magnetic plate for 5 minutes or until the bead pellet is dry. Do not overdry the bead pellet.
- Remove the tube(s) from the magnetic plate and add 16 μl water / RSB / Buffer EB to each capture reaction. Mix by pipetting until homogenized.
- Incubate at room temperature for 2 minutes
- Place the plate or tube(s) on a magnetic plate and let stand for 3 minutes or until the beads fully pellet.
- Transfer 15 μl of the clear supernatant containing the enriched library to a clean well of a 96-well thermal cycling plate, making sure not to disturb the pellet. Keep on ICE
- Validate and quantify each enriched library using:
  - Agilent Bioanalyzer High Sensitivity DNA Kit. Load 2 μl of the final sample. Average fragment length should be 375–425 bp using a range setting of 150–1,000bp.
  - Thermo Fisher Scientific Qubit dsDNA High Sensitivity Quantitation Assay.
  - Interpretation notes: Final concentration may vary and is dependent on panel size, library input, hybridization reaction size, and the number of PCR cycles Should be nice tight peak on both. Average fragment size is usually around 350bp. Distribution important for bioinformatics as this is used to estimate template length. Ideally want tight peak between 350-475. Lower quality library will have a broader peak. Can save traces to USB. <1ng/ul i'd be a little concerned but not the end of the world.
